# HCG22 as a Non-Invasive Saliva-Based lncRNA Diagnostic Marker for Oral Cancer: Evidence from an Indian Clinical Study

**DOI:** 10.1101/2025.07.30.25332447

**Authors:** Pandikannan Krishnamoorthy, Athira S Raj, Shrinkhla Singh, Akanksha Bhattacharya, Madhavan Parthasarathy, Kushal Junghare, Nilanjana Das, Ganakalyan Behera, Ashok Kumar, Vikas Gupta, Himanshu Kumar

## Abstract

**Background:** Early diagnosis of oral squamous cell carcinoma (OSCC) is limited by the lack of reliable non-invasive biomarker candidates. Long non-coding RNAs (lncRNAs) in saliva are promising non-invasive candidates due to their stability and presence across the body fluids. This study aims to discover detectable OSCC-associated lncRNAs in saliva and validate them in an Indian OSCC clinical cohort.

**Methods:** Discovery datasets comprising tissue OSCC samples (PRJNA327548, PRJNA775998, PRJNA947509, and PRJNA1031181) and healthy saliva (PRJNA985624) were analyzed using an lncRNA-focused pipeline. Robust lncRNAs consistently dysregulated in OSCC were intersected with the highly expressed lncRNAs in saliva. Clinical validation was performed through RT-PCR in tissue and saliva derived from the Indian OSCC cohort. Diagnostic potential was estimated through ROC analysis. TCGA-OSCC data were analyzed to identify prognostic relevance.

**Results:** Four robust lncRNAs were identified to be dysregulated in OSCC and have higher expression in saliva samples. Among them, HCG22 was amplified consistently in all the saliva samples and downregulated in tissue and saliva samples of OSCC patients compared to the control. Diagnostic performance was high (AUROC = 0.86 in tissues; 0.76 in saliva) in the Indian OSCC clinical cohort. Survival analysis based on TCGA-OSCC data identified a lower HCG22 association with advanced tumor stages and poorer survival.

**Conclusion:** HCG22 is a promising non-invasive salivary lncRNA with diagnostic potential for OSCC detection and prognosis.

## Introduction

Oral squamous cell carcinoma (OSCC) accounts for more than 90% of the Head and Neck cancers, the sixth most prevalent cancer worldwide. OSCC is characterized by the presence of a malignant neoplasm that originates from mucosal epithelial cells of the oral cavity with different levels of keratinization. OSCC is a major health problem in India, accounting for one-third of the global cases. As of 2022, Oral cancer is the highest reported cancer after lung cancer in India among the male population [1]. Bhopal, situated in Central India, reports the highest age-adjusted incidence rate of lip and mouth cancer among males and the second highest among females in the world [2, 3]. The progression of OSCC is multifactorial, and the known risk factors include the excessive consumption of tobacco, smokeless tobacco (paan and gutka), and HPV infections. The overall 5-year survival rate of OSCC improves up to 90 percent if it is diagnosed at an early stage [4]. Unfortunately, most of the OSCC cases are diagnosed at the later stages, and hence, the mortality rate remains high.

Tissue biopsy remains the widely utilized method of OSCC diagnosis. However, it involves expensive and highly invasive procedures that are challenging for repeated sampling and early detection at a population level. Liquid biopsy is an emerging, cost-effective, and minimally invasive method for early diagnosis that relies on utilizing the biomolecules present in the extracellular body fluids, such as blood, plasma, urine, and saliva [5]. Among them, urine and saliva are non-invasive and the most preferred samples for diagnosing any disease, including oral cancer. The dysregulation in the levels of biomarkers such as circulating cell-free tumor DNA (ct-DNA), circulating cell-free tumor RNA (cf-RNA), DNA methylation, non-coding RNAs such as long non-coding RNAs (lncRNAs), microRNAs (miRNAs), and circular RNAs (circRNAs) across different cancers and the stages in the extra-cellular fluids was reported to have excellent diagnostic potential, suitable for the affordable large-scale early diagnosis [6–8].

Among different liquid biopsies, saliva-based methods show high potential as a promising, non-invasive, and easily accessible method for OSCC diagnosis [9]. Unlike the widely utilized blood (plasma/serum) sampling, saliva collection is non-invasive, painless, and does not require specialized personnel or equipment, making it suitable for large-scale screening and repeated sampling in low-resource and rural settings. Due to its proximity to the OSCC tumor site and tumor microenvironments, the molecular biomarkers exhibit more specificity and sensitivity, in comparison to blood-based biomarkers.

LncRNAs are reported to play a crucial role in gene regulation, tumor progression, and regulation of tumor microenvironment in various cancers [10]. LncRNAs are present in body fluids, including saliva, in a stable form due to their association with exosomes or RNA-binding proteins. Due to their tumor-specific expression patterns and stability, lncRNAs are ideal biomarker candidates. However, very limited studies have attempted to identify the diagnostic potential of lncRNAs in the saliva for OSCC detection [11, 12].

In this study, by utilizing the in-house lncRNA-specific computational pipeline over the public next-generation sequencing data derived from OSCC clinical tissue samples, robust OSCC lncRNA biomarkers that have good expression in saliva were explored. The top lncRNA candidates were validated in a prospective clinical cohort derived from the tissue and saliva samples of the Indian OSCC patients to explore the diagnostic performance.

## Materials and Methods

### Study Population

The prospective clinical study was conducted at the All India Institute of Medical Sciences (AIIMS), Bhopal, between February 2024 and January 2025. The study protocol was approved by the Institutional Human Ethics Committee (AIIMS Bhopal) and the Institutional Ethical Committee (IEC) (IISER Bhopal). All the participants enrolled in the study had provided consent for the collection of clinical samples. The study participants include the patients attending the Outpatient department of ENT-HNS (Otorhinolaryngology - Head and Neck Surgery), with a history and clinical examination suggestive of oral cancer or having been operated for oral cancer, subjects with other benign oral cavity lesions, or other ENT pathologies. All the participants were subjected to routine investigations as required for their diagnosis and management. The inclusion criteria include participants aged between 20 to 80, with oral squamous cell carcinoma or a benign lesion of the oral cavity, undergoing biopsy/ definitive surgery at ENT-HNS, AIIMS Bhopal. The exclusion criteria include a previous history of treatment (surgery/radiation/chemotherapy) for OSCC, and subjects who consumed tobacco before sample collection. Only the participants who provided written consent were included in the study. The histopathological evaluation of all the samples was carried out by the oral pathologist to confirm the malignancy.

### Clinical sample collection and processing

Fresh tissue samples were collected and added to 1.5ml of RNAprotect Tissue Reagent (Qiagen) and stored at −80 °C for long-term storage. Before RNA isolation, all the tissue samples were thawed and weighed of 2mg each. Each tissue samples were resuspended in 350 μL of buffer RLT of the RNeasy Micro Kit and homogenized using the iRupt Jr homogenizer.

Unstimulated saliva samples were collected in the Paxgene Saliva collectors. The subjects were instructed not to consume any food 30 minutes before the sample collection. The samples were stored at −80 °C for long-term use. 250 μL of saliva samples were centrifuged at 12000 x g for 2 mins to remove debris, and the clear liquid portion was processed further for RNA isolation.

### RNA isolation

For isolation of RNA from the tumor samples, the RNAeasy microkit (Qiagen) was used. RNA extraction and purification were performed as per the manufacturer’s instructions. The quality of RNA and quantification were performed using the Qubit 4 Fluorometer.

For isolation of RNA from the saliva samples, the clear fluid was added with 750uL of TRIzol LS reagent and mixed well by pipetting 3-4 times. 200uL of chloroform was added and mixed properly by vortex. The samples were centrifuged at 15,000g for 15 minutes at 4 °C. The uppermost transparent layer was collected, without disturbing the intermediate layer. The samples (transparent fluid) were added with 500uL of absolute isopropanol along with 2% glycogen and incubated at 4 °C overnight. After incubation, the samples were centrifuged at 15,000g, 4 °C, for 10 mins, and a white pellet was obtained. The pellets were washed with 75% ethanol by centrifuging the samples for 5 mins at 7500g, 4 °C. The washed pellets were then air dried and then eluted in 15 μL nuclease-free water. The RNA samples were stored and maintained at −80 degrees until further use.

### cDNA synthesis and RT-PCR

The cDNA was synthesized from the tissue and saliva samples using the High-Capacity cDNA synthesis kit (Applied Biosystems), as per the manufacturer’s guidelines. Real-time PCR reactions were performed using PowerUp SYBR Green master mix (Applied Biosystems) in the QuantStudio-5 platform (Applied Biosystems). Glyceraldehyde 3-Phosphate Dehydrogenase (GAPDH) was utilized as an endogenous normalization control. The primer sequences used in this study are listed in **(Supplementary Information S4).** The RT PCR reactions were performed in triplicate, and the 2^-ΔΔCt^ method was used to calculate the fold change of each lncRNA in both tissue and saliva samples.

### Overview of the discovery datasets

To identify the lncRNAs significantly dysregulated in OSCC and potentially detectable in saliva, publicly available RNA-seq datasets were retrieved from the NCBI Sequence Read Archive (SRA). Since the goal is to identify even the novel, uncharacterized lncRNAs, only high-throughput RNA-sequencing datasets were selected. The inclusion criteria for the dataset selection include RNA-seq datasets derived from the oral tissue samples or saliva. The exclusion criteria include the datasets in which raw FASTQ files were not available publicly, in vitro cell line datasets, and datasets without any clear clinical metadata. Based on the screening, the following four OSCC tissue datasets, PRJNA327548, PRJNA775998, PRJNA947509, and PRJNA1031181, were selected for further analysis [13–15]. All these datasets included both the OSCC and Control (benign or other oral pre-malignant lesions). For the estimation of lncRNA expression in saliva, the PRJNA985624 dataset, containing RNA-seq data from saliva samples of healthy volunteers and COVID-19 patients, was used. Only healthy samples were considered.

### LncRNA-focused computational pipeline and RNA-seq analysis

The raw fastq files were downloaded from the NCBI SRA for all the datasets. The quality control, adapter trimming were performed by the fastp tool [16]. Based on the sequencing library kit utilized in the study and detected through fastp, attention was given to properly remove the adapters. The cleaned adapter-trimmed reads were pseudo-aligned to the human transcriptome reference using the Salmon tool, and the lncRNAs annotation file (gencode v47) was utilized [17]. This approach made it possible to even quantify the lncRNAs that are not well-characterized and are not included in the normal annotation files **(Fig. 1)**. The R package tximport was utilized to generate the count matrix [18]. The DESeq2 package was utilized to perform variance stabilizing normalization (vst) and differential expression analysis in the case of two groups [19]. The significant lncRNAs were identified with the threshold |logFC| >1 and adj.P.Val < 0.05. Rank-based meta-analysis was performed using the R package, RobustRankAggreg [20]. The robust lncRNAs were identified with a significance threshold of adjusted p-value (BH-method) < 0.05. The receiver operating curve analysis was performed using the R package pROC [21]. The plots were generated utilizing the R packages ggplot2, EnhancedVolcano, cowplot, and pROC.

**Figure 1:**
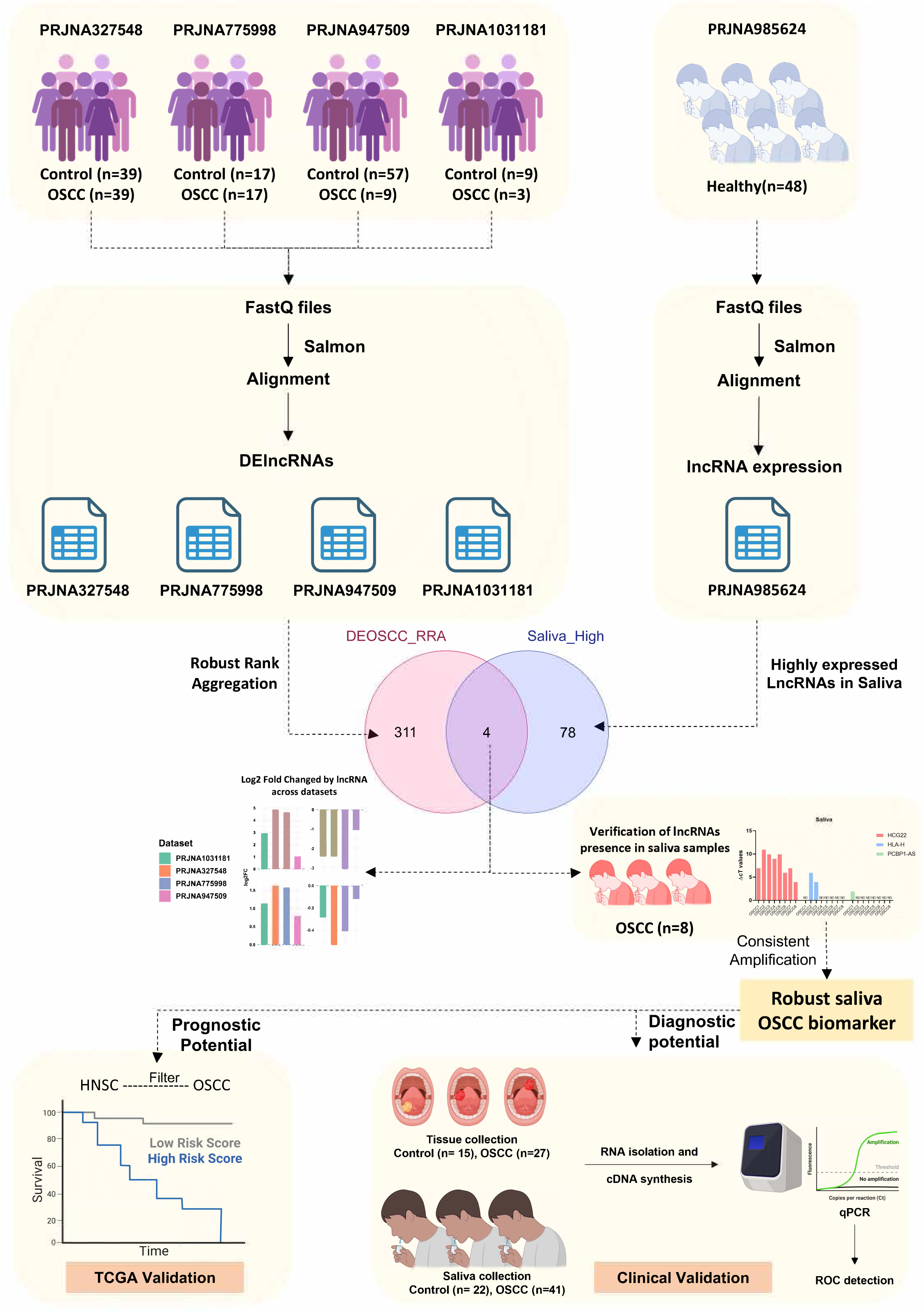
Schematic overview of the study design. Workflow for the study design to identify the significantly dysregulated lncRNA in OSCC, following a similar trend in saliva samples. Public transcriptome datasets of OSCC tissues and healthy saliva were processed through the lncRNA-focused computational pipeline. The robust lncRNAs were identified through the robust rank aggregation algorithm. The top hits were identified based on the presence in saliva and dysregulation in OSCC tissues. The top hits were further validated in a clinical cohort of Indian OSCC patients to evaluate the diagnostic potential. TCGA-OSCC datasets were utilized to evaluate the prognostic potential and risk assessment. The schematic is created with BioRender.com.

### TCGA data analysis

Since access to the raw fastq files was restricted to employ the lncRNA-specific pipeline, RPKM values were obtained from the TANRIC database [22]. The TCGAbiolinks R package was utilized to download the clinical metadata and to filter only the oral cancer-associated samples from head and neck squamous cell carcinoma (HNSC) [23]. Survival analysis was performed using the R packages survival and survminer.

### Statistical Analysis

All statistical analyses were conducted using R (v4.3.1) and GraphPad Prism (v10.0). Descriptive statistics, including medians and interquartile ranges, were used to summarize continuous variables such as age. Categorical variables, including gender and test type, were summarized using counts and percentages. Normality of data distribution was assessed using the Shapiro-Wilk test. For group comparisons, parametric tests (Student’s t-test or one-way ANOVA, Wilcoxon rank-sum test) were applied for normally distributed variables, and non-parametric tests (Mann-Whitney U test or Kruskal-Wallis test) were used for non-normally distributed variables. Chi-square or Fisher’s exact test was applied for categorical comparisons. Correlations were assessed using Pearson’s or Spearman’s correlation coefficients, as appropriate. A p-value of <0.05 was considered statistically significant.

## Results

### Identification of differentially expressed lncRNAs in OSCC tissue samples

To identify the lncRNAs associated with OSCC, high-throughput RNA sequencing datasets derived from the OSCC clinical samples were obtained from the NCBI SRA data repository. Since no dataset was available for the saliva samples, only tissue samples were considered. Four datasets, PRJNA327548, PRJNA775998, PRJNA947509, and PRJNA1031181, were selected for further analysis **(Table 1)**. The lncRNA-specific computational pipeline was used to quantify both previously characterized and novel lncRNAs using the lncRNA annotation files. The schematic of the overall study design is provided in **Fig. 1**. The segregation of the samples between the control (benign or other oral-related disease conditions) and OSCC group was visualized through the Principal component analysis (PCA) plots for the datasets PRJNA327548 **(Fig. 2A)**, PRJNA775998 **(Fig. 2B)**, PRJNA947509 **(Fig. 2C)**, and PRJNA1031181 **(Fig.2D)**, respectively. Differential expression analysis between these groups identified differentially expressed lncRNAs (DElncRNAs) with a significance threshold of |log□FC| > 1 and adjusted p-value (Padj) < 0.05. PRJNA327548 dataset comprises 39 control samples and 39 OSCC samples, and differential expression analysis identified 1127 significant lncRNAs (DElncRNAs) (denoted as red data points) and their trend is visualized through the volcano plot **(Fig. 2E)**. The global lncRNA transcriptome change was visualized through the volcano plots for the datasets PRJNA775998 **(Fig.2F)**, PRJNA947509 **(Fig.2G)**, and PRJNA1031181 **(Fig.2H)** that identified 2440 DElncRNAs, 220 DElncRNAs, and 1337 DElncRNAs, respectively **(Supplementary Information S1)**. It is also important to note that the sample size in each of the datasets is different, and the datasets, like PRJNA1031181, exhibit class imbalance **(Table 1)**.

**Figure 2:**
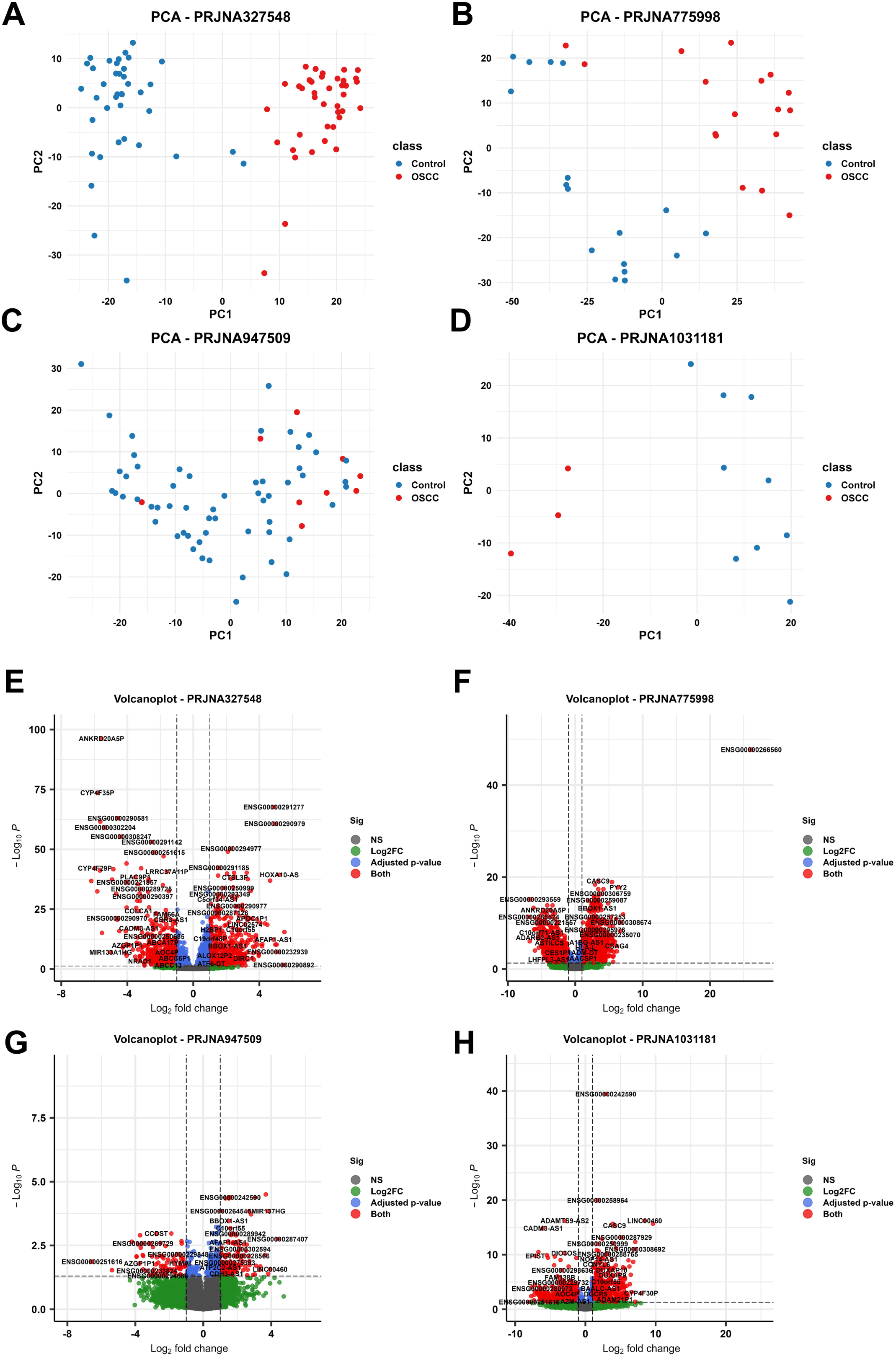
Differential expression analysis of lncRNAs in OSCC tissue datasets. (A–D) PCA plots showing the segregation of the OSCC and control samples in datasets PRJNA327548, PRJNA775998, PRJNA947509, and PRJNA1031181, respectively (E–H) Volcano plots showing differentially expressed lncRNAs (DElncRNAs) in the datasets PRJNA327548, PRJNA775998, PRJNA947509, and PRJNA1031181, respectively. Significantly dysregulated lncRNAs (|log□FC| > 1, Padj < 0.05) are highlighted in red.

**Table 1:** Public Datasets used in the study.

| **Dataset Accession** | **Country of Origin** | **Sample Type** | **Sample Size** | **Subjects / Groups** |
| --- | --- | --- | --- | --- |
| PRJNA327548 | Taiwan | Tissue | 78 | 39 OSCC,<br>39 Healthy (adjacent) |
| PRJNA775998 | China | Tissue | 34 | 17 OSCC,<br>17 Healthy (adjacent) |
| PRJNA947509 | USA | Tissue | 66 | 18 Healthy,<br>39 Premalignant lesions,<br>9 OSCC |
| PRJNA1031181 | China | Tissue | 12 | 3 OSCC,<br>3 leukoplakia (dysplasia),<br>3 leukoplakia (without<br>dysplasia),<br>3 Healthy (adjacent) |
| PRJNA985624 | USA | Saliva | 48 | 48 Control |

**Table 2.** Demographic and Clinical Characteristics of Study Participants.

| Parameter | Tissue – OSCC<br>(n = 27) | Tissue – Control<br>(n = 15) | Saliva – OSCC<br>(n = 41) | Saliva – Control<br>(n = 22) |
| --- | --- | --- | --- | --- |
| Age (years) |  |  |  |  |
| Mean ± SD | 49.7 ± 14.3 | 42.3 ± 18.6 | 50.5 ± 13.7 | 40.3 ± 14.6 |
| Median (range) | 50 (22–86) | 50 (20–71) | 52 (25–88) | 35 (20–97) |
| Sex |  |  |  |  |
| Male, n (%) | 17 (62.9%) | 14 (93.3%) | 32 (78.0%) | 16 (72.7%) |
| Female, n (%) | 10 (37.1%) | 1 (6.7%) | 9 (22.0%) | 6 (27.2%) |
| Histopathology |  |  |  |  |
| Moderately diff. SCC | 22 (81.4%) | – | 36 (87.8%) | – |
| Well diff. / Superficial SCC | 3 (11.1%) | – | 4 (9.8%) | – |
| Verrucous / Invasive SCC | 2 (7.5%) | – | 1 (2.4%) | – |
| Benign / Hyperplasia / Others | – | 15 (100%) | – | 22 (100%) |
| Subsite – OSCC |  |  |  |  |
| Lateral border of tongue | 14 (51.8%) | – | 13 (31.7%) | – |
| Buccal mucosa | 5 (18.5%) | – | 15 (36.6%) | – |
| Alveolus / Hard palate / Lip | 5 (18.5%) | – | 10 (24.4%) | – |
| Other sites | 3 (11.1%) | – | 3 (7.3%) | – |
| Clinical Staging – OSCC |  |  |  |  |
| Stage T1–T2 | 5(18.5%) | – | 7 (17.1%) | – |
| Stage T3–T4a | 20 (74.1%) | – | 27 (65.9%) | – |
| NA / Not reported | 2 (7.4%) | – | 7 (17%) | – |

SCC: Squamous Cell Carcinoma
NA: Not available or not applicable
Some cases had missing staging information.
Subsite distributions are based on available anatomic location records.

### Identification of robust OSCC-associated lncRNAs in saliva

To identify the robust lncRNAs that are significantly and consistently dysregulated and to reduce the possibility of false-positive findings due to the class imbalance, a rank-based meta-analysis algorithm was utilized. Robust rank aggregation (RRA) method relies on the ranking of the lncRNAs in each dataset rather than the group-specific approach that can be influenced by the class imbalance. After applying RRA, 315 high-confident robust OSCC-associated lncRNAs were identified **(Supplementary Information S2)**. The direction and magnitude of log fold change of the top robust lncRNAs across datasets were visualized through a horizontal bar plot **(Fig. 3A)**. Among the liquid biopsy sources, saliva is in direct contact with the OSCC tissue and tumor microenvironment, making it a promising medium for the detection of lncRNAs. The presence of lncRNAs in the saliva is understudied. The PRJNA985624 dataset comprises the RNA-seq datasets of saliva samples derived from healthy volunteers and COVID-19 patients. The raw transcriptome data of 48 healthy volunteers were subjected to lncRNA quantification using the same in-house lncRNA quantification pipeline. The expression profile of lncRNAs in saliva was summarized using the donut plot **(Fig. 3B)**. The majority of lncRNAs (74.5 %) were not detected or detected at lower levels in only a few samples. Around 18% of the total have very little expression in the majority of the samples. Only 82 lncRNAs (0.2 %) were found to be highly expressed in almost every sample consistently **(Supplementary Information S3)**. The expression of all the 82 highly expressed salivary lncRNAs, with their median values, minimal and maximal values, was visualized through the lollipop chart **(Fig. 3C)**. The next objective was to identify the robust OSCC-associated lncRNAs that are also highly expressed in saliva. A total of four lncRNAs were identified to be consistently dysregulated in OSCC tissues, identified through RRA (DEOSCC_RRA) and highly expressed in saliva (Saliva_High) **(Fig. 3D)**. Among them, ENSG00000291277 and HLA-H were consistently upregulated, while HCG22 and PCBP1-AS1 were downregulated across all four tissue datasets **(Fig. 3E)**.

**Figure 3:**
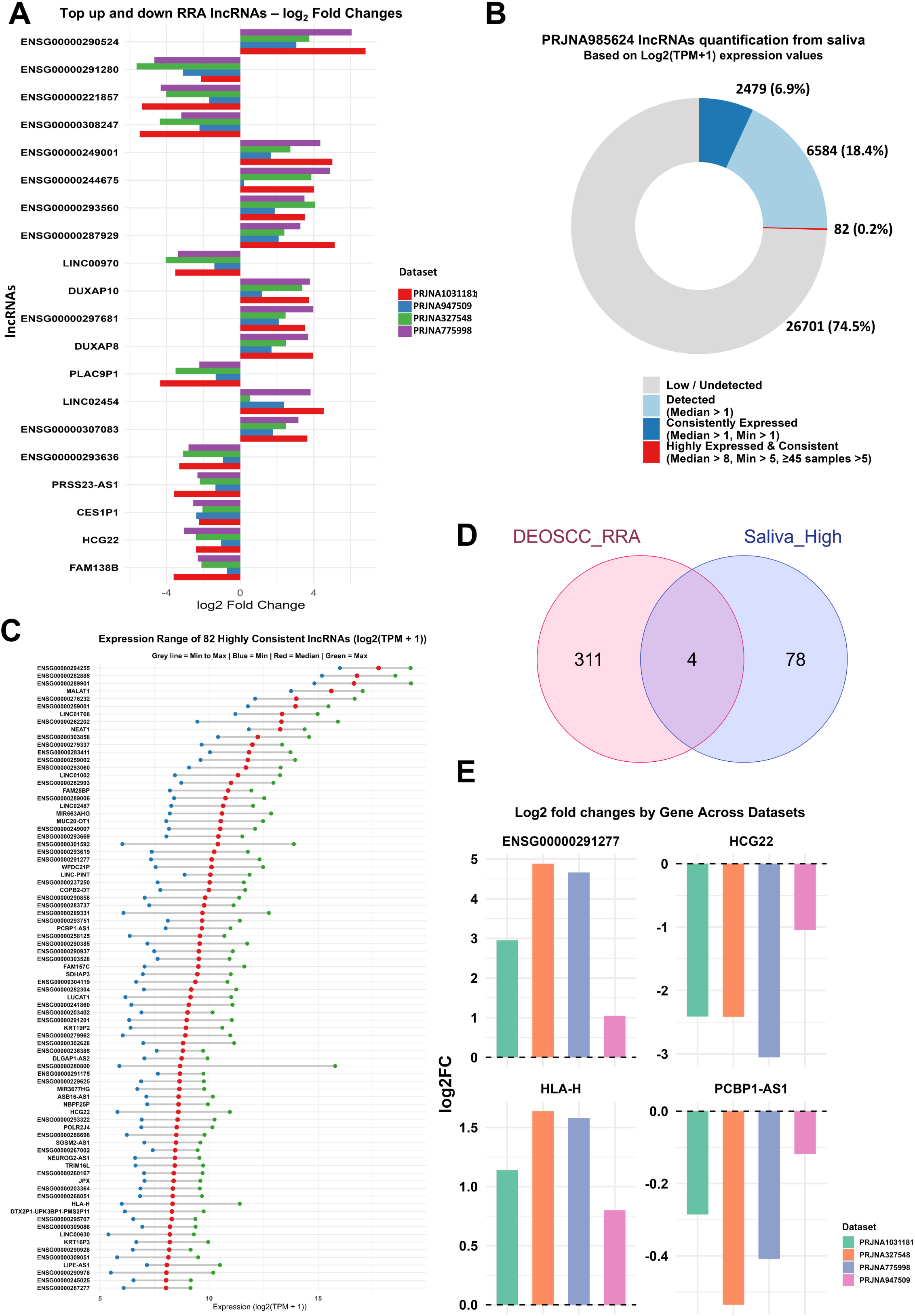
Identification of robust OSCC-associated lncRNAs and their expression in saliva. (A) Horizontal bar plot depicting the log□ fold change of top robust lncRNAs across the four tissue datasets based on the rank-based meta-analysis approach using the RobustRankAggreg package. (B) Donut chart representing the expression distribution of lncRNAs in saliva samples from healthy individuals (PRJNA985624) based on the consistent expression across all the samples and the expression level in each sample. (C) Lollipop chart showing median, minimum, and maximum expression values of the top 82 consistently highly expressed lncRNAs in saliva. Minimal values are denoted as blue, median values in red, and maximal values in green for each lncRNA. (D) Venn diagram illustrating the intersection between robust OSCC-associated lncRNAs (from RRA) and highly expressed salivary lncRNAs. Four common lncRNAs were identified. (E) Bar plots showing log□ fold changes of the four selected lncRNAs (ENSG00000291277, HLA-H, HCG22, and PCBP1-AS1) across the four OSCC tissue datasets.

### Validation of lncRNAs expression in clinical saliva samples

The expression of the four robust lncRNA candidates was further visualized across individual samples through the violin plots **(Fig. 4A)**. Among them, ENSG00000291277 had not been previously characterized and exhibits nearly 95% sequence similarity with the gene KRT17P1, making the primer design for the RT-PCR challenging. Saliva samples from eight OSCC patients were collected to validate the presence of the three lncRNAs, HCG22, HLA-H, and PCBP1-AS1, through RT-PCR using corresponding primers **(Supplementary Information S4)**. HCG22 showed high expression in all eight OSCC saliva samples, whereas HLA-H and PCBP1-AS1 were detected only in two samples and one sample, respectively **(Fig. 4B)**. Based on these results, only HCG22 was further selected for the clinical validation in a larger cohort.

**Figure 4.**
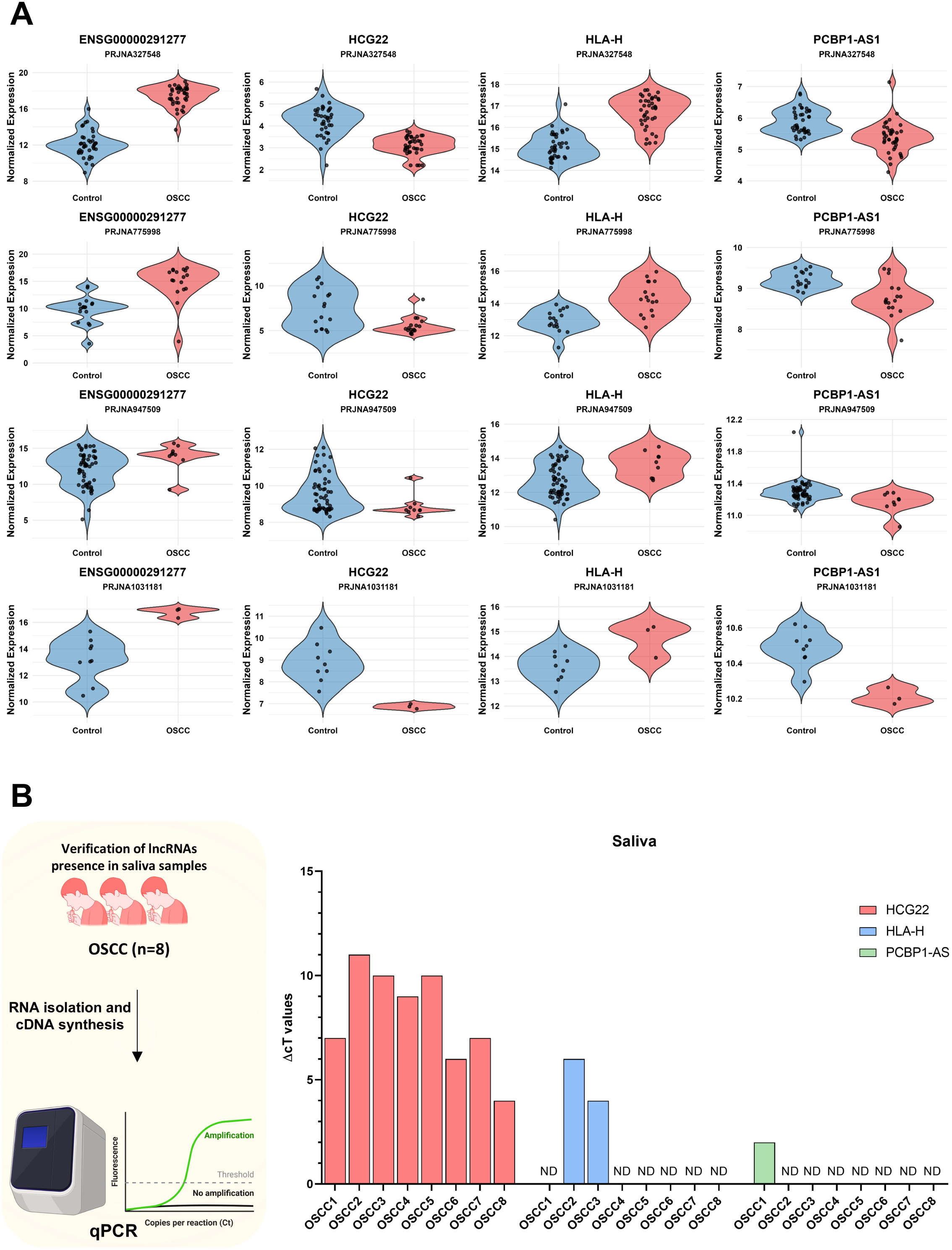
Detection of lncRNA expression in OSCC saliva samples. (A) Violin plots depicting the distribution of expression levels of the four selected lncRNAs in OSCC and control tissue samples across the discovery datasets, PRJNA327548, PRJNA775998, PRJNA947509, and PRJNA1031181, respectively. The control group is denoted in blue, whereas the OSCC group is denoted in red. (B) RT-PCR validation of HCG22, HLA-H, and PCBP1-AS1 in saliva samples from eight OSCC patients. HCG22 was consistently detected and downregulated; HLA-H and PCBP1-AS1 showed limited detection. ND denotes no detection in RT-PCR. The schematic is created with. BioRender.com

### Validation of HCG22 in public TCGA datasets

The Cancer Genome Atlas (TCGA) does not categorize oral cancer separately, but includes it under head and neck squamous cell carcinoma (HNSC). Only OSCC samples were filtered from HNSC based on the clinical metadata. The expression of HCG22 in the TCGA OSCC samples was also found to be significantly downregulated in comparison to the control group, as observed in the discovery datasets **(Fig. 5A)**. Due to the comprehensive availability of clinical metadata in the TCGA database, HCG22 dysregulated expression based on tumor grade, tumor clinical stages, and survival information was explored. No significant difference was observed in HCG22 expression based on the histological grades of OSCC samples **(Fig. 5B)**. Interestingly, the expression of HCG22 was significantly lower in advanced clinical stages **(Fig. 5C)**. Survival analysis based on the clinical metadata revealed that the lower HCG22 expression was associated with a lesser median survival time of 915 days in comparison to samples with higher HCG22 expression with median survival time of 4846 days. The results were visualized using the Kaplan-Meier curve plot **(Fig. 5D)**. Overall, HCG22 is downregulated in OSCC, and the downregulation of HCG22 expression is associated with disease progression and poor prognosis.

**Figure 5.**
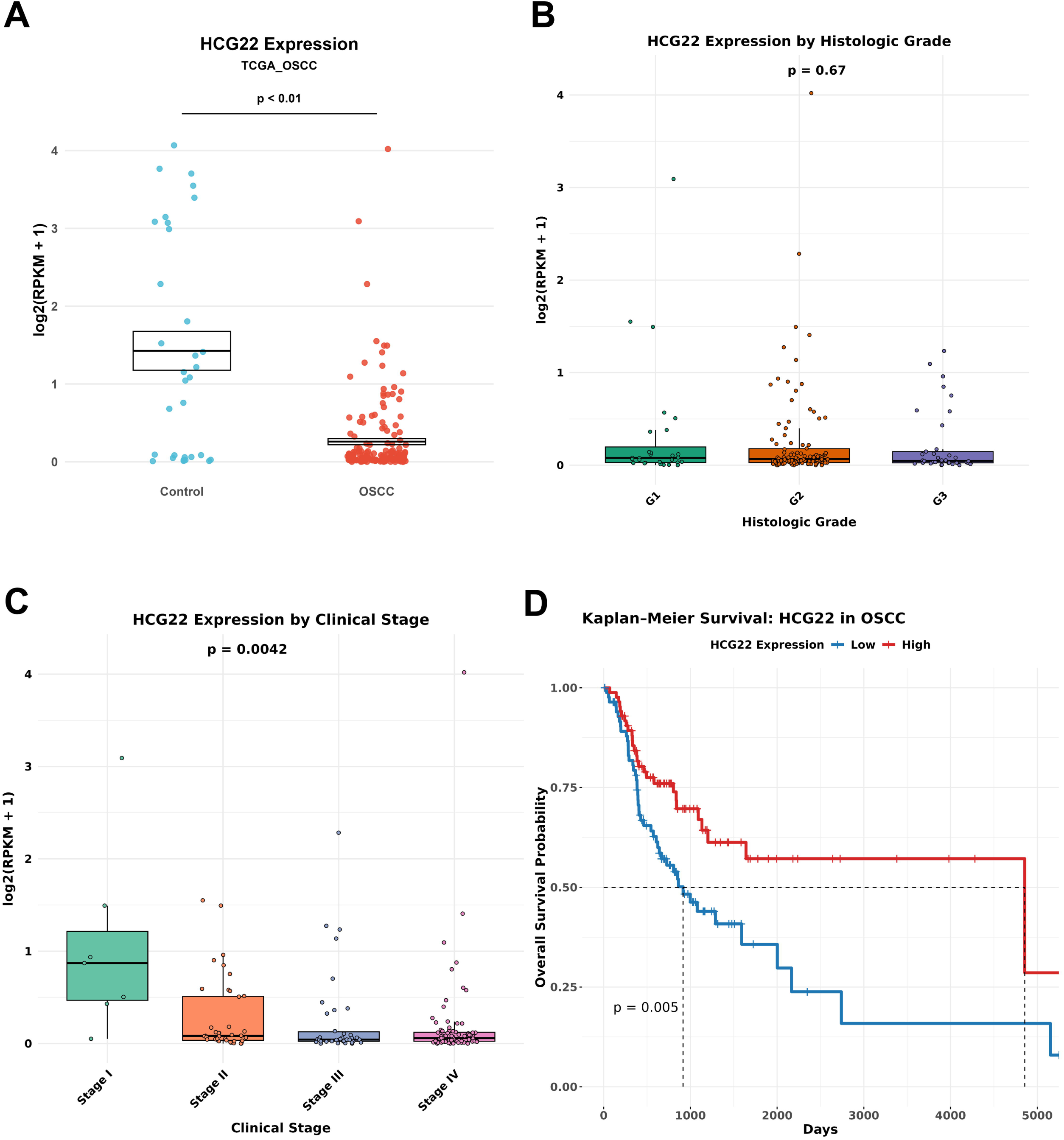
Validation of prognostic potential of HCG22 using TCGA OSCC data. (A) Box plot showing significant downregulation of HCG22 in OSCC tissue samples (n =32) compared to the control group (n = 172) in the TCGA-HNSC subset. The Wilcoxon signed-rank test was performed to identify the significance between the groups. (B) The relation between HCG22 expression and different histological grades was plotted as a bar plot. Kruskal-Wallis test was performed to identify the significance between the groups. G1: well-differentiated (Low grade), G2: Moderately differentiated (Intermediate grade), G3: Poorly differentiated (High grade). (C) Box plot depicting the significant reduction in HCG22 expression with advancing clinical stages. The Kruskal-Wallis test was performed to identify the significance between the groups. (D) Kaplan–Meier survival analysis showing reduced survival in OSCC patients with low HCG22 expression (median survival: 915 days vs. 4856 days). The red line denotes the samples with high HCG22 expression, and the Blue line represents the samples with low HCG22 expression.

### Diagnostic validation of HCG22 in Indian OSCC patients

Clinical recruitment of OSCC patients and benign controls was performed to collect the tissue and saliva samples for RT-PCR-based quantification of HCG22 **(Fig. 6A)**. The expression of HCG22 was downregulated in the tissue of OSCC patients in comparison to the benign controls **(Fig. 6B)**. The ROC curve analysis revealed higher diagnostic accuracy with a ROC value of 0.86 **(Fig. 6C)**. HCG22 expression was higher in the saliva samples of healthy volunteers based on the computational analysis and was also consistently expressed in eight saliva samples of OSCC patients. Hence, we hypothesized that HCG22 may also be downregulated in the saliva of OSCC patients, in comparison to the control subjects. Salivary RNA was isolated from the OSCC patients and controls, and RT-PCR-based quantification of HCG22 was performed. The downregulation of HCG22 was confirmed in the saliva of OSCC patients in comparison to the control group **(Fig. 6D)**. Further, ROC analysis yielded an AUC of 0.76, revealing the good diagnostic potential in saliva samples **(Fig. 6E)**.

**Figure 6.**
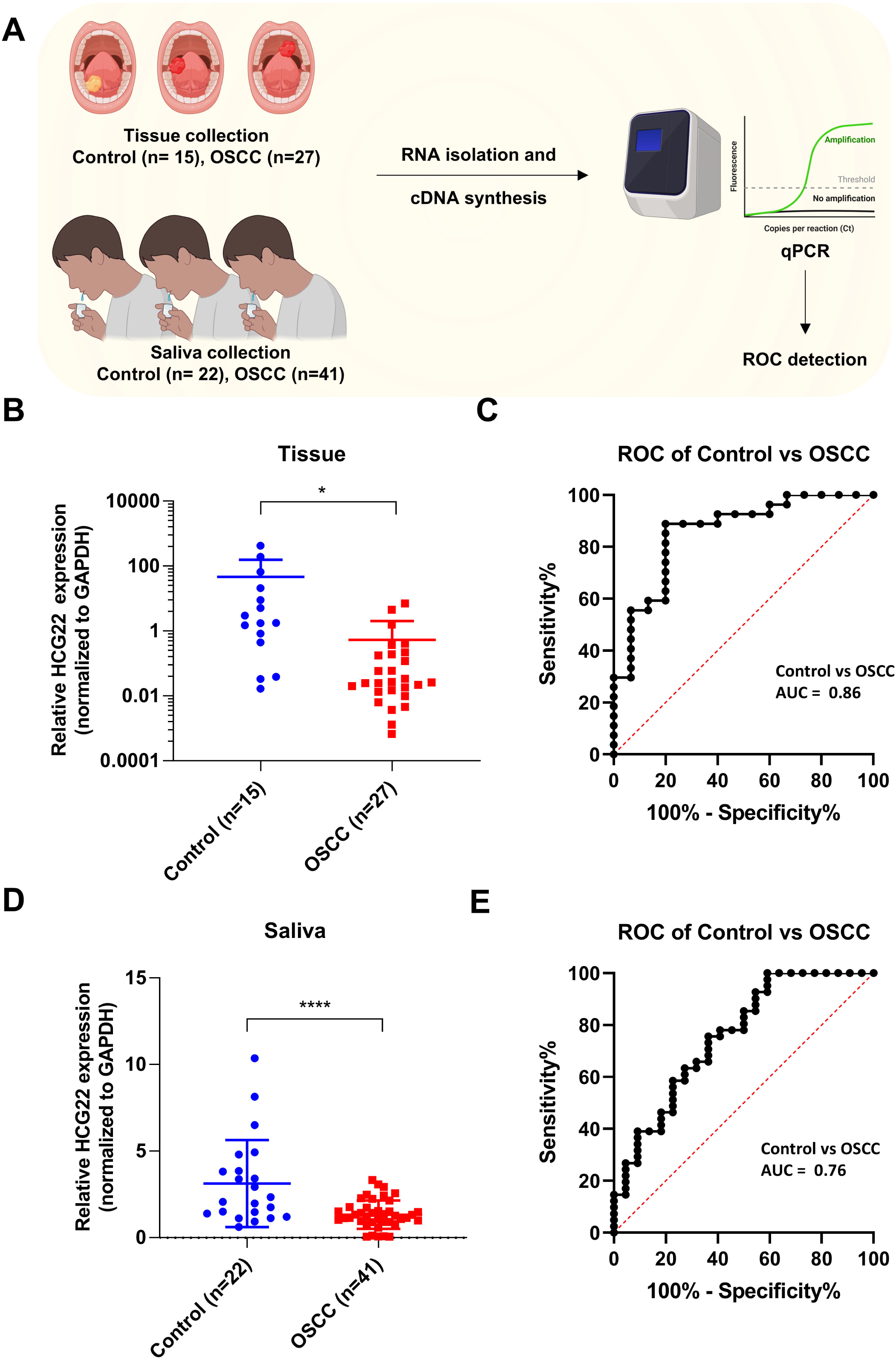
Diagnostic validation of HCG22 in clinical OSCC tissue and saliva samples. (A) Schematic overview of clinical sample collection and experimental workflow. The schematic is created with. BioRender.com. (B) RT-PCR validation of HCG22 expression in OSCC tissue compared to benign controls in the Indian clinical cohort. Data are means from technical triplicate values. Two-tailed unpaired t test was performed to identify the significance between the groups ∗∗∗∗, p < 0.0001, ∗∗∗p < 0.001, ∗∗p < 0.01, and ∗p < 0.05. (C) ROC curve analysis showing diagnostic performance of HCG22 in tissue (AUC = 0.86). (D) RT-PCR validation of HCG22 expression in OSCC saliva compared to benign controls in the Indian clinical cohort. Data are means from technical triplicate values. Two-tailed unpaired t test was performed to identify the significance between the groups ∗∗∗∗, p < 0.0001, ∗∗∗p < 0.001, ∗∗p < 0.01, and ∗p < 0.05. (E) ROC curve for HCG22 expression in saliva (AUC = 0.76), indicating diagnostic potential for non-invasive detection.

## Discussion

Early diagnosis of OSCC can improve the survival outcomes, and hence, there is an immediate need for a non-invasive, cost-effective diagnosis method suitable for large-scale screening. Saliva-based liquid biopsy offers higher diagnostic potential for OSCC due to its proximity to the tumor site and microenvironment. lncRNAs were reported to be present in the body fluids such as saliva, plasma, and urine with good stability and have higher diagnostic potential.

Limited studies explored the diagnostic potential of lncRNAs in the saliva of OSCC patients. In a study, six cancer-associated lncRNAs based on the existing reports were chosen, and their expression in the saliva of 9 OSCC patients was quantified [11]. Among them, MALAT1 expression was detected in all the saliva samples. A recent study identified upregulation of MALAT1 in the saliva of 20 OSCC patients [12]. Non-tumor control was not utilized in this study. Another study reported the reduced expression of salivary XIST lncRNA to be associated with the increased risk of OSCC [24]. However, this lncRNA expression is biased towards female patients but not detected in men. Another study identified LINC00657 to be upregulated in the saliva of 12 OSCC patients in comparison to the control group [25]. All these studies measured the pre-determined lncRNA based on the existing reports and validated its presence in a limited sample size. Moreover, lncRNAs that are highly dysregulated in the OSCC tissues compared to the control samples and follow a similar pattern in saliva have higher diagnostic and therapeutic relevance.

To overcome all the limitations of the existing studies, this study aims to identify the lncRNAs of higher diagnostic potential in the saliva of OSCC patients and validate the diagnostic potential in the Indian OSCC patients with enough sample size **(Fig. 1)**. By utilizing the lncRNA-focused computational pipeline, this study follows a systematic approach that includes the identification of robust lncRNAs from the public clinical transcriptome datasets derived from OSCC tissue samples and the identification of highly expressed salivary lncRNAs in healthy subjects. The identified hits were validated in both the tissue and saliva samples of the Indian OSCC clinical cohort.

Due to the lack of high-throughput RNA-sequencing data derived from the saliva of OSCC patients, the initial objective was to identify the consistently dysregulated robust lncRNAs dysregulated in OSCC patients, in comparison with the control group. Four independent clinical RNA-seq datasets were analyzed using a lncRNA-specific computational pipeline, allowing for the quantification of both known and novel lncRNAs. Despite the varied sample sizes and class imbalance among the datasets, the rank-based meta-analysis approach allowed us to identify robust, significant OSCC-associated lncRNAs **(Fig. 3A)**. This approach was valuable in integrating heterogeneous transcriptome datasets, reducing the dataset-specific biases that can lead to the identification of biologically meaningful hits.

The dataset PRJNA985624 was subjected to the same lncRNA-specific pipeline to identify the top lncRNAs present in the saliva samples of healthy subjects **(Fig. 3B)**. As expected, only 82 lncRNAs (0.2% of the total) were highly expressed in almost all the 48 saliva samples. By intersecting these 82 lncRNAs with the robust OSCC-associated lncRNAs identified in the tissue datasets, four lncRNAs were identified with good expression in saliva and significantly dysregulated in OSCC tissues.

The detected lncRNAs from the computational pipeline were validated in the saliva samples of Indian OSCC patients. Only HCG22 expression was consistently amplified in the saliva of all 8 patients **(Fig. 4B)**. Further, HCG22 expression was downregulated in the OSCC patients in the TCGA dataset. The reduced expression was associated with the advanced clinical stages of OSCC, and the patients with lower HCG22 expression exhibit shorter median survival time, revealing its prognostic potential **(Fig. 5D)**.

The clinical recruitment of OSCC patients was performed to validate the diagnostic potential of HCG22. The expression of HCG22 was observed to discriminate between OSCC and the control group in both the tissue and saliva samples with high diagnostic accuracy (AUC = 0.86 and 0.76, respectively), highlighting the translational relevance (**Fig. 6)**.

To our knowledge, this is the first study that identified and validated lncRNAs in the saliva of OSCC patients with a higher sample size. HCG22 was previously reported to play a tumor suppressive role in OSCC by downregulating miR-425-5p expression [26]. Another study identified the anti-tumor property of HCG22 through the inhibition of Akt, mTOR, and Wnt/B-catenin pathways [27]. The downregulation of HCG22 in the saliva of OSCC patients was observed in this study, and the finding that the lower expression of HCG22 is associated with shorter survival time reveals the diagnostic, prognostic, and translational potential of HCG22 in OSCC diagnosis.

A few limitations of the study include the need for a higher sample size with enough clinical stage information and survival information in the clinical cohort. Although ENSG00000291277 was identified as one of the top hits, due to the challenges in primers with specific amplification, this lncRNA was not validated in clinical samples. Future long-read sequencing and advanced primer design strategies may enable better characterization of such unannotated transcripts.

Multi-center clinical studies with higher sample sizes are needed to further explore the diagnostic potential of HCG22 in OSCC diagnosis, prognosis, and risk stratification in real-world applications.

## Conclusion

Overall, through the integrated approach of computational analysis of multiple independent clinical datasets and clinical validation in Indian OSCC patients, HCG22 emerged as a robust downregulated lncRNA in OSCC tissue and saliva samples. HCG22 expression was correlated with the disease stage and poor survival outcome in the TCGA dataset. The stable detection in all the saliva samples and the downregulation in OSCC saliva samples demonstrate the strong diagnostic performance, particularly in non-invasive saliva samples. These findings support HCG22 as a promising non-invasive saliva-based biomarker candidate for OSCC detection and prognosis.

## Supplementary Information legends

**Supplementary Information 1:** Differentially expressed lncRNAs in each of the discovery datasets, PRJNA327548, PRJNA775998, PRJNA947509, and PRJNA1031181.

**Supplementary Information 2:** Significant robust lncRNAs consistently dysregulated in all four discovery tissue datasets identified through the robust rank aggregation method with a significance threshold of adj.P-value < 0.05.

**Supplementary Information 3:** List of lncRNAs with high and consistent expression in 48 saliva samples with their minimum, median, and maximum values.

**Supplementary Information 4:** List of the primers and oligonucleotides used in the study for amplification using RT-PCR.

## Data availability

All the data have been presented in the main text and supplementary materials. Any further details can be requested by contacting the corresponding author.

## Acknowledgement

We acknowledge all the authors who have deposited the raw transcriptome data in the public databases. The authors thank IISERB, Indian Council of Medical Research (ICMR), India, for the funding and for access to the computational workstation.

## Contributions

PK, ASR, and SS drafted the manuscript. PK, HK performed the conceptualization and design of the study. PK performed all the computational analysis. VG, GB, and AK acquired the clinical samples after ethical approval. AB, MP, KJ, and ND performed sample annotation, collection, and maintained clinical information. PK, ASR, and SS performed all the sample processing, RNA isolation, and expression quantification. HK and VG acquired the funds for the project. HK supervised the study design. HK and AK reviewed and edited the manuscript.

## Ethics declarations

### Ethics approval and consent to participate

This clinical study was approved by the Institutional Human Ethics Committee of AIIMS Bhopal (IHEC-LOP/2024/EL0129) and the Institutional Ethical Committee of IISER Bhopal (IISERB/IEC/2024-I/01). All the samples were collected only after informed consent from the clinical subjects.

### Consent for publication

All the authors have read and approved the manuscript.

## References

1. Sathishkumar, K., et al., Cancer incidence estimates for 2022 & projection for 2025: Result from National Cancer Registry Programme, India. Indian J Med Res, 2022. 156(4&5): p. 598–607.

2. Sharma, S., et al., Oral cancer statistics in India on the basis of first report of 29 population-based cancer registries. J Oral Maxillofac Pathol, 2018. 22(1): p. 18–26.

3. Gupta, B., A. Ariyawardana, and N.W. Johnson, Oral cancer in India continues in epidemic proportions: evidence base and policy initiatives. Int Dent J, 2013. 63(1): p. 12–25.

4. Yang, J., et al., Survival analysis of age-related oral squamous cell carcinoma: a population study based on SEER. Eur J Med Res, 2023. 28(1): p. 413.

5. Ma, L., et al., Liquid biopsy in cancer current: status, challenges and future prospects. Signal Transduct Target Ther, 2024. 9(1): p. 336.

6. Krishnamoorthy, P., et al., Global Pan-cancer serum miRNA classifier across 13 cancer types: Analysis of 46,349 clinical samples. medRxiv, 2025: p. 2025.02.07.25321821.

7. Xu, C., et al., A Circulating Panel of circRNA Biomarkers for the Noninvasive and Early Detection of Pancreatic Ductal Adenocarcinoma. Gastroenterology, 2024. 166(1): p. 178–190 e16.

8. Li, Y., et al., Pan-cancer characterization of immune-related lncRNAs identifies potential oncogenic biomarkers. Nat Commun, 2020. 11(1): p. 1000.

9. Kumar, P., S. Gupta, and B.C. Das, Saliva as a potential non-invasive liquid biopsy for early and easy diagnosis/prognosis of head and neck cancer. Transl Oncol, 2024. 40: p. 101827.

10. Beylerli, O., et al., Long noncoding RNAs as promising biomarkers in cancer. Noncoding RNA Res, 2022. 7(2): p. 66–70.

11. Tang, H., et al., Salivary lncRNA as a potential marker for oral squamous cell carcinoma diagnosis. Mol Med Rep, 2013. 7(3): p. 761–6.

12. Shalaby, R., et al., MALAT1 as a potential salivary biomarker in oral squamous cell carcinoma through targeting miRNA-124. Oral Dis, 2024. 30(4): p. 2075–2083.

13. Chen, T.W., et al., APOBEC3A is an oral cancer prognostic biomarker in Taiwanese carriers of an APOBEC deletion polymorphism. Nat Commun, 2017. 8(1): p. 465.

14. Zhao, W., et al., Unidirectional alteration of methylation and hydroxymethylation at the promoters and differential gene expression in oral squamous cell carcinoma. Front Genet, 2023. 14: p. 1269084.

15. Khan, M.M., et al., Total RNA sequencing reveals gene expression and microbial alterations shared by oral pre-malignant lesions and cancer. Hum Genomics, 2023. 17(1): p. 72.

16. Chen, S., et al., fastp: an ultra-fast all-in-one FASTQ preprocessor. Bioinformatics, 2018. 34(17): p. i884–i890.

17. Patro, R., et al., Salmon provides fast and bias-aware quantification of transcript expression. Nat Methods, 2017. 14(4): p. 417–419.

18. Soneson, C., M.I. Love, and M.D. Robinson, Differential analyses for RNA-seq: transcript-level estimates improve gene-level inferences. F1000Res, 2015. 4: p. 1521.

19. Love, M.I., W. Huber, and S. Anders, Moderated estimation of fold change and dispersion for RNA-seq data with DESeq2. Genome Biol, 2014. 15(12): p. 550.

20. Kolde, R., et al., Robust rank aggregation for gene list integration and meta-analysis. Bioinformatics, 2012. 28(4): p. 573–80.

21. Robin, X., et al., pROC: an open-source package for R and S+ to analyze and compare ROC curves. BMC Bioinformatics, 2011. 12: p. 77.

22. Li, J., et al., TANRIC: An Interactive Open Platform to Explore the Function of lncRNAs in Cancer. Cancer Res, 2015. 75(18): p. 3728–37.

23. Colaprico, A., et al., TCGAbiolinks: an R/Bioconductor package for integrative analysis of TCGA data. Nucleic Acids Res, 2016. 44(8): p. e71.

24. Shieh, T.M., et al., Lack of Salivary Long Non-Coding RNA XIST Expression Is Associated with Increased Risk of Oral Squamous Cell Carcinoma: A Cross-Sectional Study. J Clin Med, 2021. 10(19).

25. Tarrad, N.A.F., et al., "Salivary LINC00657 and miRNA-106a as diagnostic biomarkers for oral squamous cell carcinoma, an observational diagnostic study". BMC Oral Health, 2023. 23(1): p. 994.

26. Fu, Y., et al., Long non-coding RNA HCG22 inhibits the proliferation, invasion and migration of oral squamous cell carcinoma cells by downregulating miR-425-5p expression. Exp Ther Med, 2022. 23(3): p. 246.

27. Wang, M., et al., Assessment of multiple pathways involved in the inhibitory effect of HCG22 on oral squamous cell carcinoma progression. Mol Cell Biochem, 2021. 476(6): p. 2561–2571.

